# Hemodynamic Mechanisms Underlying Orthostatic Hypotension in Stroke Survivors: A Cross-Sectional Study

**DOI:** 10.1101/2024.12.18.24319275

**Authors:** Kazuaki Oyake, Ayumi Mochida, Masakiyo Terashi, Mahiro Hasegawa, Akari Saito, Kunitsugu Kondo, Yohei Otaka, Kimito Momose

**Affiliations:** Department of Physical Therapy, School of Health Sciences, Shinshu University, Nagano, Japan; Department of Rehabilitation Medicine, Tokyo Bay Rehabilitation Hospital, Chiba, Japan; ‘Present address’ Department of Physical Therapy, School of Rehabilitation Sciences, Health Sciences University of Hokkaido, Japan; Department of Rehabilitation Medicine, Fujita Health University School of Medicine, Aichi, Japan

**Keywords:** cardiac output, cerebrovascular disease, rehabilitation, tilt-table test, total peripheral resistance

## Abstract

**Background:** Orthostatic hypotension is an important consideration for stroke survivors, given its association with adverse outcomes, such as cardiovascular diseases and falls. Understanding the hemodynamic mechanisms underlying orthostatic hypotension is essential for selecting the appropriate treatment based on individual hemodynamic patterns. However, the relative contribution of changes in cardiac output and total peripheral resistance to orthostatic hypotension remains unclear in stroke survivors.

**Objective:** To determine whether orthostatic hypotension is more strongly associated with a marked decrease in cardiac output or an impaired increase in total peripheral resistance among individuals with stroke.

**Methods:** This cross-sectional study included 23 individuals with stroke (13 males, mean [SD] age 63.7 [12.1] years, mean time since stroke 85.1 [34.1] days) recruited from an intensive inpatient rehabilitation ward between June 2022 and November 2023. Participants underwent a head-up tilt test to assess orthostatic hypotension and associated changes in cardiac and total peripheral resistance indices. The head-up tilt test protocol consisted of a 5-min period in the supine position followed by a 5-min period with a 70° head-up tilt. Orthostatic hypotension was defined as a reduction in systolic blood pressure of at least 20 mmHg or diastolic blood pressure of at least 10 mmHg during the test.

**Results:** Orthostatic hypotension was identified in five participants (22%). During the head-up tilt test, these individuals demonstrated a significantly greater increase in the cardiac index (p = 0.023) and decrease in the total peripheral resistance index (p = 0.002) than those without orthostatic hypotension.

**Conclusions:** Our results suggest that an impaired increase in total peripheral resistance upon standing mainly contributes to orthostatic hypotension in individuals with stroke. These findings advance the understanding of the hemodynamic mechanisms underlying orthostatic hypotension in the stroke population and may guide the implementation of targeted therapeutic strategies.

**Highlights:**

- Orthostatic hypotension is associated with a higher risk of adverse outcomes.
- Mechanisms of orthostatic hypotension were examined in stroke survivors.
- Orthostatic hypotension may be associated with impaired vasoconstriction.
- Findings may aid in managing orthostatic hypotension during stroke rehabilitation.

## Introduction

Stroke is one of the primary causes of disability, with an increased risk of recurrence [1], cardiovascular diseases [2], dementia [3], and falls [4]. Orthostatic hypotension is also associated with a higher risk of these adverse outcomes [5-8], with a prevalence of 13.0–52.1% among stroke survivors [9-14]. Appropriate rehabilitation interventions to prevent or treat orthostatic hypotension may reduce future risks in this population.

Orthostatic hypotension occurs due to a marked decrease in cardiac output and/or an impaired increase in total peripheral resistance upon standing [15]. The marked decrease in cardiac output reflects an excessive reduction in venous return, while the impaired increase in total peripheral resistance is associated with inadequate baroreflex-mediated compensatory vasoconstriction [15]. Treatment strategies differ based on the underlying mechanism. For instance, compression garments or fludrocortisone are potential treatments for marked cardiac output decrease, while midodrine may be prescribed for an impaired increase in total peripheral resistance [15]. Thus, understanding the hemodynamic mechanisms underlying orthostatic hypotension is essential for selecting the appropriate treatment based on individual hemodynamic patterns. However, to the best of our knowledge, no studies have investigated the relative contributions of cardiac output and total peripheral resistance changes to orthostatic hypotension in stroke survivors.

We aimed to examine whether orthostatic hypotension is more strongly associated with a marked decrease in cardiac output or an impaired increase in total peripheral resistance in individuals with stroke. In this population, orthostatic hypotension may result from complex interactions between various conditions such as baroreflex dysfunction, muscle weakness, hypovolemia, and comorbidities [5, 12, 13, 16, 17]. Older adults who share similar potential causes of orthostatic hypotension with stroke survivors have been reported to show a decrease in total peripheral resistance upon standing [18, 19]. Thus, we hypothesized that the orthostatic hypotension would be more strongly associated with an impaired increase in total peripheral resistance than with changes in cardiac output in individuals with stroke.

## Material and Methods

### Study design

This study used a cross-sectional, observational design. The ethics committee of Shinshu University approved the study protocol on May 12, 2022 (approval number: 5514). All participants provided written informed consent before enrollment in the study. This study adhered to the principles of the Declaration of Helsinki of 1964, as revised in 2013.

### Participants

We recruited participants from an intensive inpatient rehabilitation ward between June 2022 and November 2023. The inclusion criteria were as follows: (1) age 40–90 years; (2) stroke onset within 180 days; and (3) ability to cooperate with study procedures. The exclusion criteria were as follows: (1) limited range of motion and/or pain affecting the head-up tilt test; (2) moderate to severe dementia as defined by a Mini-Mental State Examination score of < 21 [20, 21]; (3) implanted metal devices such as a pacemaker and joint arthroplasty; (4) bilateral hemiparetic stroke; (5) unstable medical conditions, such as arrhythmias and uncontrolled diabetes mellitus; and (6) any comorbid neurological disorders. Demographic and clinical data, including age, sex, stroke type, and comorbidities, were extracted from medical records.

### Data collection

All assessments were performed in a quiet room at a comfortable temperature between 16:00 and 18:00. Participants were instructed to refrain from eating and consuming caffeinated products for at least 2 h and to avoid vigorous exercise for at least 12 h before the assessments [22].

### Head-up tilt test

The participants rested in a supine position on a motorized tilt table (SPR-7001, SAKAI Medical Co., Ltd., Tokyo, Japan) for 10 min to establish stable baseline hemodynamics [22]. We then measured hemodynamic variables during a 5-min supine period, followed by a 5-min passive head-up tilt to 70° [22]. The test was promptly terminated if a participant experienced severe symptoms, such as presyncope.

Throughout the head-up tilt test, the participants were instructed to regulate their breathing at a rate of 0.25 Hz (approximately 15 breaths per minute) using a computer metronome with audio cues. This procedure minimized the influence of respiratory fluctuations on heart rate variability assessments [23].

At the end of the test, participants were instructed to report the severity of orthostatic symptoms, such as dizziness and palpitations, experienced during the 5-min upright posture using a visual analog scale. Participants marked a 100-mm horizontal line with a pen, where 0 mm indicated no symptoms and 100 mm represented syncope or presyncope [24, 25].

### Assessments of hemodynamic and heart rate variability responses to head-up tilt

We measured hemodynamic variables using a non-invasive impedance cardiography device (Task Force Monitor 3040i, CNSystems Medizintechnik GmbH., Graz, Austria). A digital cuff was placed on the middle finger of the non-paretic hand for continuous arterial blood pressure measurement. The non-paretic arm was supported by a sling to maintain the finger cuff at the level of the heart. Beat-to-beat systolic blood pressure, diastolic blood pressure, and mean arterial pressure were obtained using the vascular unloading technique [26] and automatically corrected to oscillometric blood pressure values from the paretic arm.

Heart rate, R-R intervals, stroke volume, cardiac output, and total peripheral resistance were also measured on a beat-to-beat basis. Heart rate and R-R intervals were derived from six-lead electrocardiography. Stroke volume was calculated using an improved transthoracic impedance cardiography method [27], with three short-band electrodes placed on the participants: one on the neck and two below the thorax. Stroke volume was calculated using the following formula:

Stroke volume = Vth × LVET × (dZ/dt)max/Z0,

where Vth is the electrical participating thoracic volume, LVET is the left ventricular ejection time, (dZ/dt)max is the maximal rate of impedance reduction for a given heartbeat, and Z0 is the base impedance. Cardiac output was calculated as the product of the heart rate and stroke volume. Total peripheral resistance was derived using Ohm’s law:

Total peripheral resistance = (mean arterial pressure/cardiac output) × 80.

We normalized stroke volume, cardiac output, and total peripheral resistance to the estimated body surface area to derive the stroke index, cardiac index, and total peripheral resistance index (TPRI), respectively. We excluded data acquired during postural changes to avoid motion artifacts. Data on systolic and diastolic blood pressure, mean arterial pressure, heart rate, stroke index, cardiac index, and TPRI were averaged every minute. Supine values were defined as the average values obtained from the final minute of the supine period. Orthostatic hypotension was defined as a reduction in systolic blood pressure of at least 20 mmHg or diastolic blood pressure of at least 10 mmHg during the test [28, 29].

Heart rate variability analysis was performed to assess cardiac autonomic function. Heart rate variability parameters for the supine position were determined from the R-R intervals recorded during the 5-min supine period, while those for the upright position were derived from the 5-min upright period. We applied a threshold-based artifact correction algorithm with a medium filter to remove ectopic beats and artifacts [25, 30]. This filter identified R-R intervals that deviated by more than 0.25 s from the local average. The identified artifacts were replaced with interpolated values using cubic spline interpolation. These calculations were performed using the Kubios HRV standard software version 3.5 (Kubios Oy, Kuopio, Finland).

We calculated the frequency-domain variables of heart rate variability using fast Fourier transform analysis, which used a Welch periodogram method with a window width of 256 s and 50% window overlap. Low-frequency (0.04–0.15 Hz) and high-frequency (0.15–0.40 Hz) spectral components were collected as absolute values of power. The low-frequency component of heart rate variability represents a combination of sympathetic and parasympathetic activity and baroreflex function, whereas the high-frequency component of heart rate variability primarily reflects vagal tone [23, 31]. Furthermore, the low-to-high frequency ratio of heart rate variability was used as an indicator of the sympathetic-parasympathetic balance, with a higher ratio reflecting greater sympathetic activity [23, 31].

### Assessment of body composition

We measured body composition using a multi-frequency bioelectrical impedance analyzer (InBody S10; InBody Co., Ltd., Seoul, South Korea) with the participants in a supine position. Eight electrodes were attached to the thumbs, middle fingers, and ankles of both sides.

We collected data on the skeletal muscle mass index, total body water, and fat-free mass. The skeletal muscle mass index was calculated as the sum of the skeletal muscle mass in the upper and lower extremities divided by the height squared. We defined low muscle mass as a skeletal muscle mass index of < 7.0 kg/m2 for male individuals and < 5.7 kg/m2 for female individuals [32]. In addition, we calculated the ratio of total-body water to fat-free mass, as a lower ratio indicates hypovolemia induced by bed rest [33].

### Assessment of functional outcomes

We assessed motor function and independence in performing activities of daily living as functional outcomes. To evaluate motor impairments in the paretic upper and lower extremities, we used the motor items of the Stroke Impairment Assessment Set [34]. The degree of independence in activities of daily living was measured using the Functional Independence Measure [35].

### Statistical analyses

We calculated the required sample size using G*power version 3.1.9.2 (Heinrich-Heine-Universität, Düsseldorf, Germany). Based on previous studies comparing hemodynamic responses between middle-aged and older adults with and without orthostatic hypotension [18, 19], we assumed an effect size of 0.90 with an alpha level of 0.05 and a power of 0.80 for independent t-tests. Given the expected prevalence of orthostatic hypotension of 20% in stroke survivors [14], the required sample size was calculated to be 50 participants (10 with orthostatic hypotension and 40 without orthostatic hypotension).

The normality of distribution for all continuous variables was assessed using the Shapiro–Wilk test. Heart rate variability parameters showed significant deviation from normality (p < 0.05) and were logarithmically transformed prior to analysis. Characteristics of participants were compared between two groups (with and without orthostatic hypotension) using the unpaired t-test for continuous variables, the Mann–Whitney U test for ordinal variables, and Fisher’s exact test for dichotomous variables. To test our hypothesis, hemodynamic responses to the head-up tilt test were compared between the groups using two-way repeated-measures analysis of variance (ANOVA) with the group as the between-subject factor and time (supine and 1–5 minutes post-standing) as the within-subject factor. In addition, comparisons of heart rate variability responses to the head-up tilt test between the groups were analyzed using a two-way repeated-measures ANOVA with the group as the between-subject factor and position (supine and upright positions) as the within-subject factor.

Based on our hypothesis that orthostatic hypotension in stroke survivors would be more strongly associated with an impaired increase in total peripheral resistance, we further investigated factors associated with orthostatic TPRI changes. We examined the associations between the orthostatic change in TPRI and the characteristics of participants, hemodynamic variables and heart rate variability parameters in the supine position, and the orthostatic changes in hemodynamic variables and heart rate variability parameters. These associations were analyzed using Pearson’s product-moment correlation coefficient, the unpaired t-test, and Spearman’s rank correlation coefficient based on variable types. Orthostatic changes in hemodynamic variables were calculated by subtracting the measurements in the supine position from those at the lowest TPRI during the upright period. Statistical analyses were performed using GraphPad Prism version 9.00 for Windows (GraphPad Software, San Diego, California, United States). Statistical significance was set at a p < 0.05.

## Results

### Participants

Fig. 1 presents the flow of participant enrollment. Of the 177 individuals with stroke assessed for eligibility, 154 were excluded, primarily because of moderate to severe dementia (n = 97). Owing to an unexpected equipment malfunction that could not be repaired within the study timeframe, only 23 participants were included in the analysis instead of the planned 50 participants. Table 1 presents the characteristics of the participants. All participants were in the subacute phase of stroke recovery [36] and participated in approximately 180 min of intensive daily rehabilitation.

**Figure 1.**
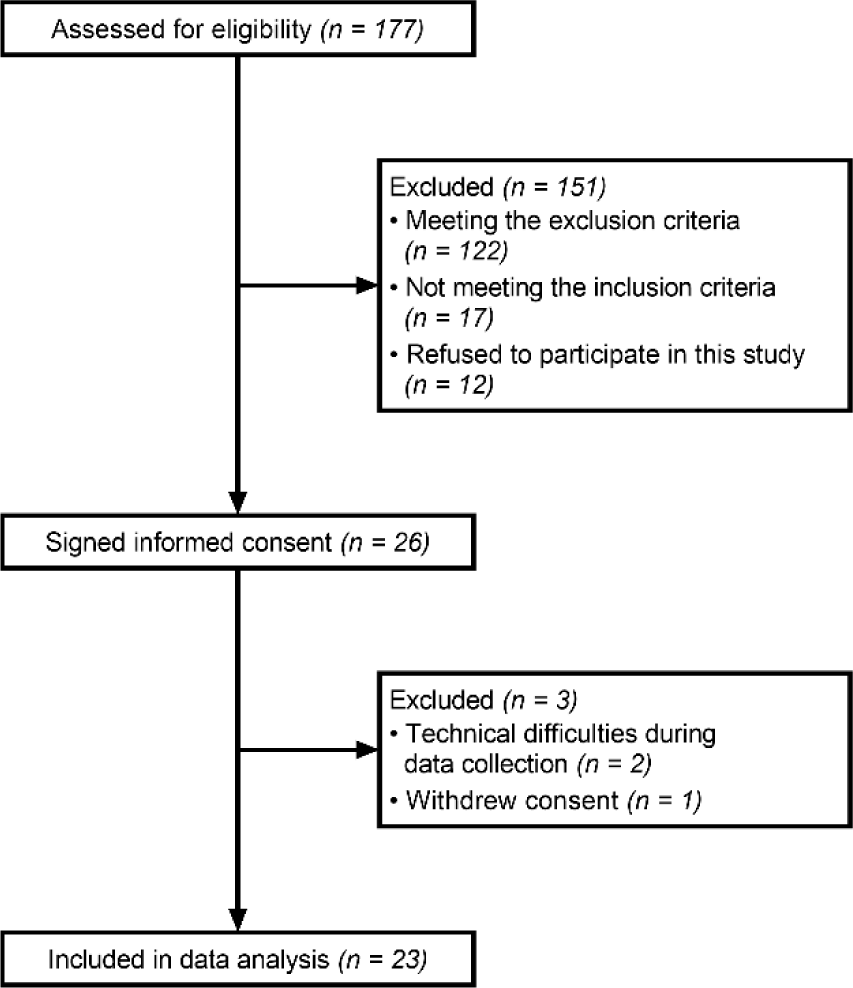
Flow diagram of participant enrollment.

**Table 1.** Characteristics of participants.

| Variable | Overall (*n* = 23) | With OH (*n* = 5) | Without OH (*n* = 18) | p-value |
| --- | --- | --- | --- | --- |
| Age, years | 63.7 (12.1) | 64.6 (17.0) | 63.5 (11.0) | 0.862 |
| Sex, male/female | 13/10 | 4/1 | 9/9 | 0.339 |
| Height, m | 1.63 (0.08) | 1.66 (0.07) | 1.62 (0.08) | 0.447 |
| Weight, kg | 66.6 (13.9) | 61.2 (10.7) | 67.3 (14.7) | 0.405 |
| Body mass index, kg/m<sup>2</sup> | 24.8 (4.9) | 22.2 (2.8) | 25.5 (5.1) | 0.189 |
| Stroke type, ischemia/hemorrhage | 15/8 | 3/2 | 12/6 | 0.999 |
| Side of motor paresis, right/left | 8/15 | 1/4 | 7/11 | 0.621 |
| Time since stroke, days | 85.1 (34.1) | 82.0 (33.8) | 85.9 (35.2) | 0.825 |
| Antihypertensive medications |  |  |  |  |
| Calcium channel blockers | 13 | 2 | 11 | 0.618 |
| Angiotensin II receptor blockers | 4 | 1 | 3 | 0.999 |
| Diuretics | 4 | 0 | 4 | 0.539 |
| Beta-blocker | 2 | 1 | 1 | 0.395 |
| Comorbidities |  |  |  |  |
| Hypertension | 19 | 3 | 16 | 0.194 |
| Diabetes mellitus | 3 | 0 | 4 | 0.539 |
| Skeletal muscle mass index, kg/m<sup>2</sup> | 7.58 (1.07) | 7.42 (1.37) | 7.62 (1.02) | 0.724 |
| Low skeletal muscle mass, male/female | 3/0 | 2 | 1 | 0.107 |
| Ratio of total body-water to fat-free mass | 0.741 (0.003) | 0.712 (0.003) | 0.746 (0.025) | 0.690 |
| Stroke Impairment Assessment Set motor score, points | 16 (10–20) | 10 (7–20) | 17 (13–20) | 0.599 |
| Functional Independence Measure motor score, points | 68 (54–78) | 52 (49–73) | 69 (58–79) | 0.421 |
| Functional Independence Measure cognitive score, points | 31 (29–34) | 31 (31–33) | 31 (29–34) | 0.966 |

Values are presented as the mean (SD), median (1st quartile–3rd quartile), or number. P-values indicate significant differences between the groups with and without OH.
OH, orthostatic hypotension

All the participants completed the head-up tilt test without any adverse orthostatic symptoms. The median (1st quartile–3rd quartile) visual analog scale score was 0 (0–8) mm. However, five participants (22%) met the criteria for orthostatic hypotension in this study. Two participants met both the systolic and diastolic blood pressure criteria, while three exhibited only diastolic orthostatic hypotension. The characteristics of participants did not differ significantly between the groups with and without orthostatic hypotension (p > 0.05, Table 1).

### Hemodynamic and heart rate variability responses to the head-up tilt test

Fig. 2 shows the hemodynamic and heart rate variability responses to the head-up tilt test in both groups with and without orthostatic hypotension. Table 2 summarizes the changes in hemodynamic variables from supine to each minute of the 5-min upright period, stratified by orthostatic hypotension status.

**Figure 2.**
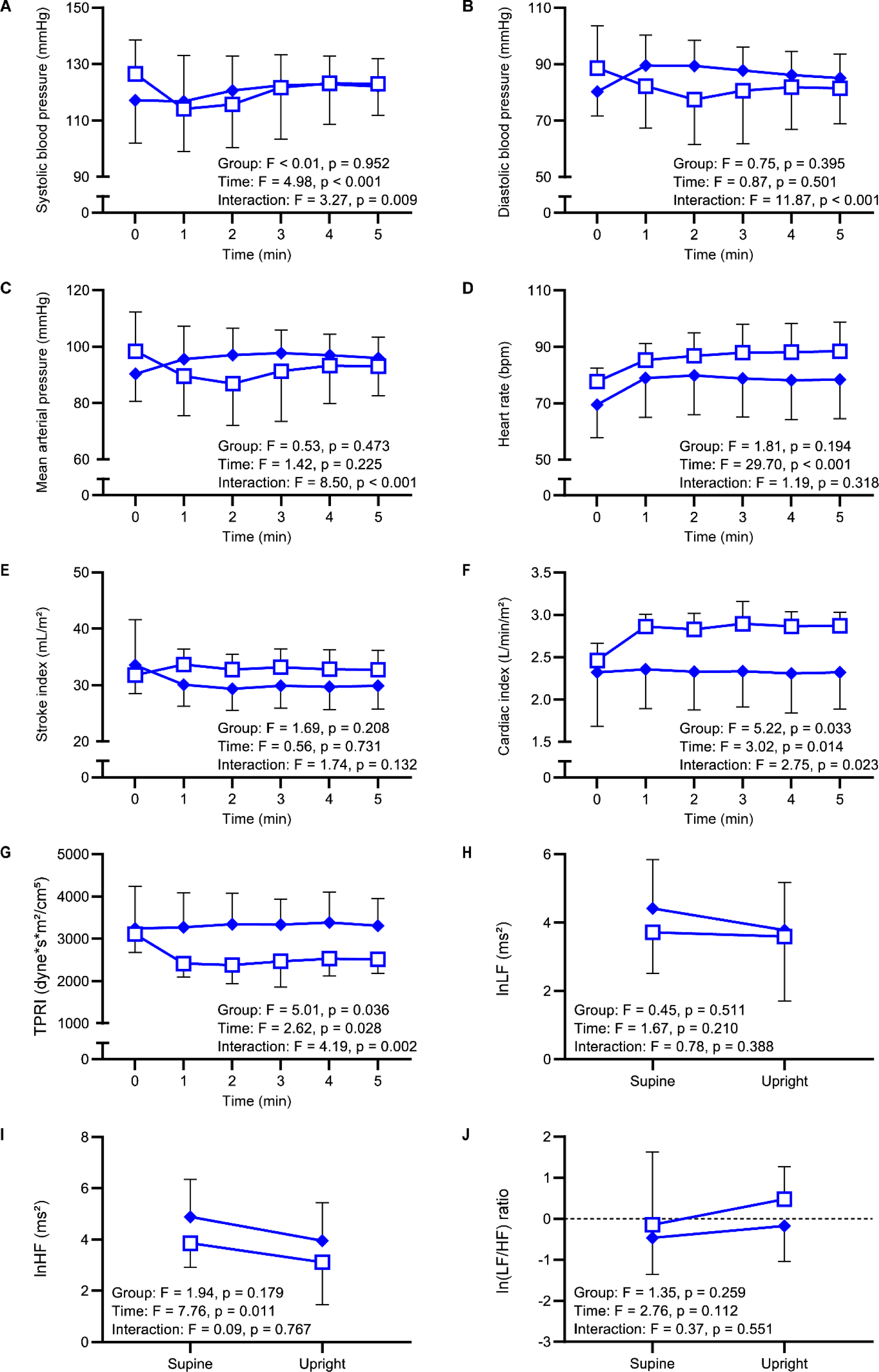
Comparisons of the hemodynamic and heart rate variability responses to the head-up tilt test between the groups with and without orthostatic hypotension. Hemodynamic variables include the (A) systolic blood pressure, (B) diastolic blood pressure, (C) mean arterial pressure, (D) heart rate, (E) stroke index, (F) cardiac index, and (G) TPRI. Heart rate variability parameters are the (H) lnLF, (I) lnHF, and (J) ln(LF/HF) ratio. The white squares and the blue diamonds represent the mean values in the groups with and without orthostatic hypotension, respectively. The error bars indicate the SD. Data at 0 on the x-axis corresponds to data at the final 1 min of the supine period. lnHF, logarithmic high-frequency component of heart rate variability; lnLF, logarithmic low-frequency component of heart rate variability; ln(LF/HF) ratio, logarithmic low-to-high frequency ratio of heart rate variability; TPRI, total peripheral resistance index.

**Table 2.** Changes in hemodynamic variables from the supine position at each minute during the 5-min upright period categorized by the presence of orthostatic hypotension.

| Variable | Group | Change from the supine position |  |  |  |  |
| --- | --- | --- | --- | --- | --- | --- |
|  |  | 1 min | 2 min | 3 min | 4 min | 5 min |
| Systolic blood pressure, mmHg | With OH | -12.5 (6.6) | -10.9 (9.7) | -4.8 (10.3) | -3.3 (7.3) | -3.5 (4.8) |
|  | Without OH | -0.3 (3.48) | 3.5 (9.3) | 5.3 (11.1) | 5.7 (9.5) | 5.0 (8.8) |
| Diastolic blood pressure, mmHg | With OH | -6.4 (3.1) | -11.2 (3.2) | -8.0 (5.2) | -6.8 (4.0) | -7.1 (6.6) |
|  | Without OH | 9.3 (8.3) | 9.2 (6.5) | 7.6 (5.7) | 5.9 (5.8) | 4.8 (4.6) |
| Mean arterial pressure, mmHg | With OH | -8.9 (4.2) | -11.6 (4.4) | -7.2 (6.6) | -5.2 (5.0) | -5.4 (5.5) |
|  | Without OH | 5.2 (8.4) | 6.7 (6.6) | 7.4 (7.4) | 6.7 (7.1) | 5.7 (5.7) |
| Heart rate, bpm | With OH | 7.6 (2.6) | 9.0 (9.7) | 10.2 (6.8) | 10.4 (6.4) | 10.7 (6.6) |
|  | Without OH | 9.5 (6.2) | 10.4 (6.2) | 9.3 (5.8) | 8.7 (6.2) | 8.9 (5.6) |
| Stroke index, mL/m<sup>2</sup> | With OH | 1.88 (3.39) | 0.98 (3.72) | 1.37 (4.04) | 1.02 (3.78) | 0.97 (3.61) |
|  | Without OH | -3.4 (7.5) | -4.2 (8.0) | -3.6 (8.4) | -3.8 (7.8) | -3.6 (7.5) |
| Cardiac index, L/min/m<sup>2</sup> | With OH | 0.40 (0.25) | 0.37 (0.35) | 0.43 (0.43) | 0.40 (0.35) | 0.41 (0.34) |
|  | Without OH | 0.04 (0.45) | 0.01 (0.48) | 0.01 (0.49) | -0.01 (0.46) | 0.00 (0.43) |
| TPRI, dyne*s*m<sup>2</sup>/cm<sup>5</sup> | With OH | -696.2 (324.7) | -732.7 (402.4) | -645.6 (594.6) | -582.0 (505.7) | -595.3 (462.8) |
|  | Without OH | 31.5 (632.8) | 106.2 (611.6) | 93.2 (688.2) | 143.2 (605.5) | 68.0 (674.8) |

Values represent the mean (SD) of the changes in hemodynamic variables from the supine position.
OH, orthostatic hypotension; TPRI, total peripheral resistance index.

The two-way repeated-measures ANOVA revealed significant interactions between group and time for systolic blood pressure (F_(5,105)_ = 3.27, p = 0.009; Fig. 2A), diastolic blood pressure (F_(5,105)_ = 11.87, p < 0.001; Fig. 2B), and mean arterial pressure (F_(5,105)_ = 8.50, p < 0.001; Fig. 2C). After standing, the group without orthostatic hypotension showed increasing trends in these variables, whereas the group with orthostatic hypotension demonstrated decreasing trends (Table 2).

For heart rate, although neither the group × time interaction (F_(5,105)_ = 1.19, p = 0.318) nor the main effect of the group (F_(1,21)_ = 1.81, p = 0.194) was significant, there was a significant main effect of time (F_(5,105)_ = 29.70, p < 0.001; Fig. 2D), with both groups showing increased values after standing (Table 2). The stroke index showed neither significant group × time interaction (F_(5,105)_ = 1.74, p = 0.132) nor a main effect of the group (F_(1,21)_ = 1.69, p = 0.208; Fig. 2E).

Analysis of the cardiac index (Fig. 2F) and TPRI (Fig. 2G) revealed significant group × time interactions (cardiac index: F_(5,105)_ = 2.75, p = 0.023; TPRI: F_(5,105)_ = 4.19, p = 0.002) and main effects of the group (cardiac index: F_(1,21)_ = 5.22, p = 0.033; TPRI: F_(1,21)_ = 5.01, p = 0.036). These results indicate that compared to the group without orthostatic hypotension, the group with orthostatic hypotension demonstrated significantly greater increases in the cardiac index and more pronounced decreases in the TPRI after standing (Table 2). In all participants with orthostatic hypotension, a higher cardiac index and a lower TPRI remained throughout the upright period than in the supine period.

For the logarithmic low-frequency component (lnLF), the logarithmic high-frequency component (lnHF), and the logarithmic low-to-high frequency ratio (ln(LF/HF) ratio), there were neither significant group × time interactions (lnLF: F_(1,21)_ = 0.78, p = 0.388; lnHF: F_(1,21)_ = 0.09, p = 0.767; ln(LF/HF) ratio: F_(1,21)_ = 0.37, p = 0.551) nor main effects of group (lnLF: F_(1,21)_ = 0.45, p = 0.511; lnHF: F_(1,21)_ = 1.94, p = 0.179; ln(LF/HF) ratio: F_(1,21)_ = 1.35, p = 0.259).

### Factors associated with an impaired orthostatic increase in TPRI

No significant associations were found between the orthostatic change in TPRI and the characteristics of participants (p > 0.05; Table 3). Correlations of orthostatic change in TPRI with hemodynamic variables and heart rate variability parameters during the head-up tilt test are shown in Fig. 3 and Table 4. We found that a greater orthostatic decrease in TPRI was associated with lower supine values of the stroke index (r = 0.550, 95% confidence interval [CI] = 0.178–0.784, p = 0.006; Fig. 3A), the cardiac index (r = 0.570, 95% CI = 0.208–0.796, p = 0.004; Fig. 3B), and a higher supine TPRI (r = -0.613, 95% CI = -0.818 to -0.269, p = 0.002; Fig. 3C). No other significant correlations were found between the orthostatic change in TPRI and supine hemodynamic variables or heart rate variability parameters (p > 0.05; Table 4).

**Figure 3.**
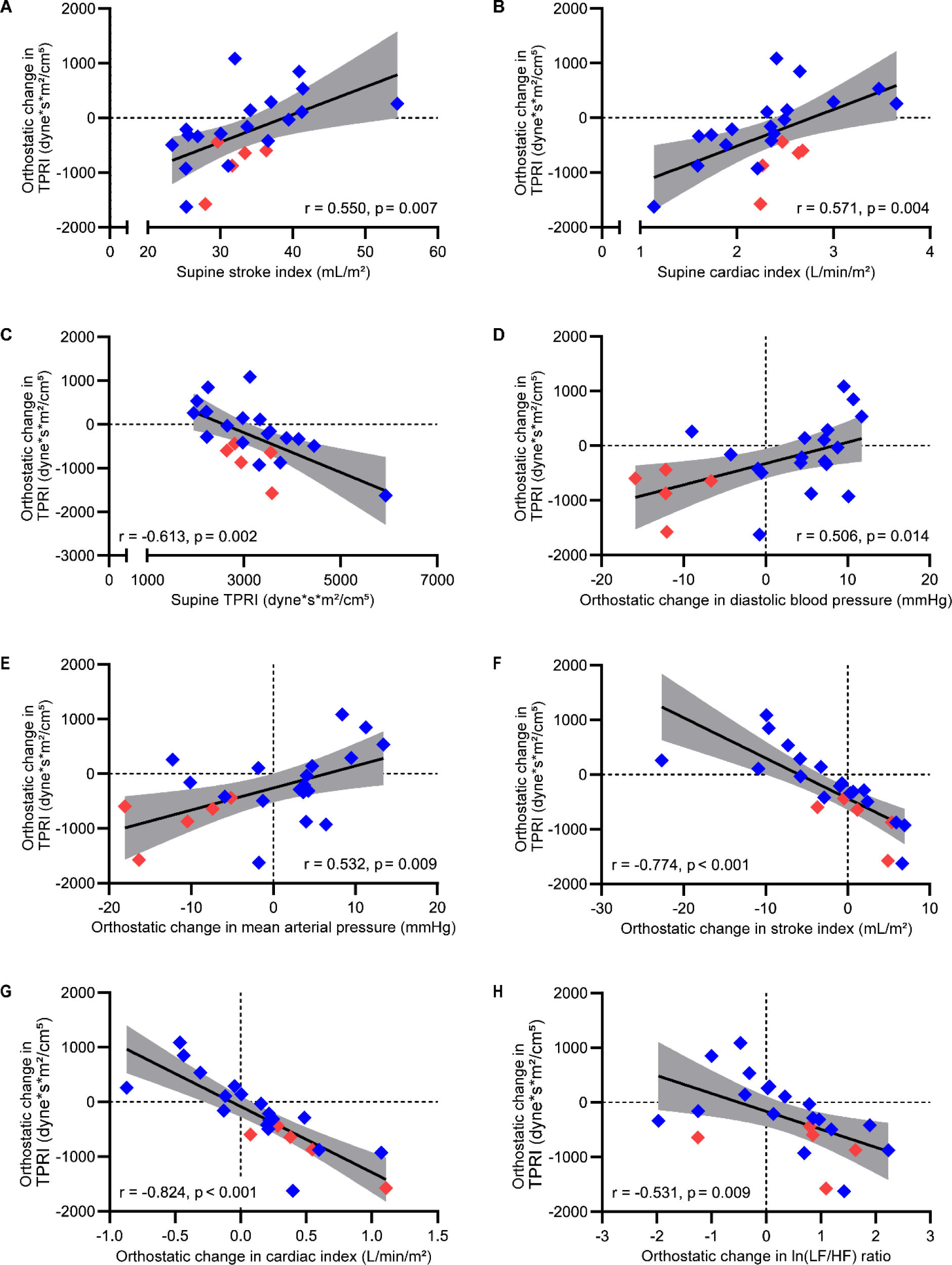
Correlations of orthostatic change in the total peripheral resistance index with hemodynamic variables and heart rate variability parameters during the head-up tilt test. Scatter plots showing correlations between orthostatic change in TPRI and (A) supine stroke index, (B) supine cardiac index, (C) supine TPRI, (D) orthostatic change in diastolic blood pressure, (E) orthostatic change in mean arterial pressure, (F) orthostatic change in the stroke index, (G) orthostatic change in the cardiac index, and (H) orthostatic change in the ln(LF/HF) ratio. The red and blue diamonds represent participants with and without orthostatic hypotension, respectively. The solid black line represents the regression line, while the shaded area indicates the 95% confidence interval of the regression line. Vertical dashed lines in panels D-H indicate no change from supine to the upright position. Horizontal dashed lines indicate no change in TPRI from supine to the upright position. ln(LF/HF) ratio, logarithmic low-to-high frequency ratio of heart rate variability; TPRI, total peripheral resistance index.

**Table 3.** Associations between orthostatic change in total peripheral resistance index and the characteristics of participants.

| Variable | Orthostatic change in TPRI |  |
| --- | --- | --- |
|  | Statistics | p-value |
| Age | $r = -0.089$ | 0.687 |
| Sex | $t = 1.631$ | 0.118 |
| Height | $r = -0.224$ | 0.305 |
| Weight | $r = -0.214$ | 0.328 |
| Body mass index | $r = -0.131$ | 0.552 |
| Stroke type | $t = -0.647$ | 0.524 |
| Side of motor paresis | $t = 0.475$ | 0.639 |
| Time since stroke | $r = -0.230$ | 0.291 |
| Antihypertensive medications |  |  |
| Calcium channel blockers | $t = 0.406$ | 0.689 |
| Angiotensin II receptor blockers | $t = 1.452$ | 0.161 |
| Diuretics | $t = 1.589$ | 0.127 |
| Beta-blocker | $t = 1.950$ | 0.065 |
| Comorbidities |  |  |
| Hypertension | $t = -1.331$ | 0.198 |
| Diabetes mellitus | $t = 1.319$ | 0.201 |
| Skeletal muscle mass index | $r = -0.277$ | 0.202 |
| Low muscle mass | $t = 1.002$ | 0.328 |
| Ratio of total body-water to fat-free mass | $r = -0.274$ | 0.206 |
| Stroke Impairment Assessment Set motor score | $\rho = -0.227$ | 0.297 |
| Functional Independence Measure motor score | $\rho = -0.071$ | 0.748 |
| Functional Independence Measure cognitive score | $\rho = -0.306$ | 0.156 |

TPRI, total peripheral resistance index.

**Table 4.** Correlations of orthostatic change in total peripheral resistance index with hemodynamic variables and heart rate variability parameters during the head-up tilt test.

| Variable | Orthostatic change in TPRI |  |  |
| --- | --- | --- | --- |
|  | r | 95% CI | p-value |
| Supine systolic blood pressure | -0.086 | -0.481 to 0.338 | 0.683 |
| Supine diastolic blood pressure | -0.090 | -0.484 to 0.335 | 0.683 |
| Supine mean arterial pressure | -0.120 | -0.507 to 0.307 | 0.584 |
| Supine heart rate | 0.147 | -0.283 to 0.527 | 0.504 |
| Supine stroke index | **0.550** | **0.178–0.784** | **0.006** |
| Supine cardiac index | **0.571** | **0.208–0.796** | **0.004** |
| Supine TPRI | **-0.613** | **-0.818 to -0.269** | **0.002** |
| Supine lnLF | 0.249 | -0.182 to 0.600 | 0.252 |
| Supine lnHF | 0.155 | -0.274 to 0.533 | 0.479 |
| Supine ln(LF/HF) ratio | 0.114 | -0.313 to 0.502 | 0.606 |
| Orthostatic change in systolic blood pressure | 0.343 | -0.081 to 0.662 | 0.109 |
| Orthostatic change in diastolic blood pressure | **0.506** | **0.119–0.760** | **0.014** |
| Orthostatic change in mean arterial pressure | **0.532** | **0.154–0.775** | **0.009** |
| Orthostatic change in heart rate | 0.077 | -0.346 to 0.474 | 0.767 |
| Orthostatic change in stroke index | **-0.774** | **-0.899 to -0.530** | **<0.001** |
| Orthostatic change in cardiac index | **-0.824** | **-0.923 to -0.624** | **<0.001** |
| Orthostatic change in lnLF | -0.272 | -0.615 to 0.158 | 0.210 |
| Orthostatic change in lnHF | 0.223 | -0.208 to 0.582 | 0.306 |
| Orthostatic change in ln(LF/HF) ratio | **-0.531** | **-0.774 to -0.152** | **0.009** |

Values marked in bold indicate a significant correlation.
CI, confidence interval; lnHF, logarithmic high-frequency component of heart rate variability; ln(LF/HF) ratio, logarithmic low-to-high frequency ratio of heart rate variability; lnLF, logarithmic low-frequency component of heart rate variability; r, Pearson's product-moment correlation coefficient; TPRI, total peripheral resistance index.

Regarding orthostatic changes, a greater decrease in TPRI was significantly correlated with larger reductions in diastolic blood pressure (r = 0.506, 95% CI = 0.119– 0.760, p = 0.014; Fig. 3D) and mean arterial pressure (r = 0.532, 95% CI = 0.154–0.775, p = 0.009; Fig. 3E). Additionally, a greater TPRI decrease was associated with larger increases in the stroke index (r = -0.774, 95% CI = -0.899 to -0.530, p < 0.001; Fig. 3F), cardiac index (r = -0.824, 95% CI = -0.923 to -0.624, p < 0.001; Fig. 3G), and ln(LF/HF) ratio (r = -0.531, 95% CI = -0.774 to -0.152, p = 0.009; Fig. 3H). No other significant correlations were found between the orthostatic change in TPRI and orthostatic changes in the remaining parameters (p > 0.05; Table 4).

## Discussion

To the best of our knowledge, this study is the first to examine the hemodynamic mechanisms underlying orthostatic hypotension in stroke survivors. Compared to the group without orthostatic hypotension, the group with orthostatic hypotension demonstrated greater increases in the cardiac index and larger decreases in TPRI during the head-up tilt test. These results support our hypothesis that orthostatic hypotension may be more strongly associated with an impaired increase in total peripheral resistance than with changes in cardiac output in individuals with stroke. Our findings may provide clinicians with valuable insights into managing orthostatic hypotension during stroke rehabilitation.

The prevalence of orthostatic hypotension in this study population (22%) was similar to data from our previous study of inpatients with subacute stroke (24%) [14]. In addition, all participants remained asymptomatic during the head-tilt test, regardless of the presence or absence of orthostatic hypotension. A study also reported that many individuals with orthostatic hypotension were asymptomatic despite substantial reductions in systolic blood pressure and low upright systolic blood pressure [37]. These findings highlight the importance of measuring orthostatic changes in blood pressure rather than depending on symptoms alone to indicate orthostatic hypotension. Although habituation and cerebral autoregulation are possible explanations for the absence of orthostatic symptoms, further research is needed to elucidate the mechanisms underlying this impaired symptom recognition [37].

The transition from the supine to the upright position causes blood pooling in the lower extremities and splanchnic veins, which may lead to reduced venous return, stroke volume, and cardiac output. In healthy adults, baroreflex-mediated compensation increases heart rate, cardiac contractility, and total peripheral resistance, ultimately raising the mean arterial pressure [29, 38]. Studies have reported statistically significant increases in blood pressure variables, heart rate, and total peripheral resistance, along with decreases in stroke volume and cardiac output, during this postural change in healthy individuals [39-41]. In contrast, our participants with orthostatic hypotension showed sustained higher cardiac index and lower TPRI values relative to the supine values throughout the upright period. These results suggest that inadequate baroreflex-mediated vasoconstriction mainly contributes to orthostatic hypotension in individuals with stroke. Additionally, we found the associations of orthostatic decrease in TPRI with orthostatic increases in the stroke index, cardiac index, and ln(LF/HF) ratio as well as with orthostatic reductions in diastolic blood pressure and mean arterial pressure. These results suggest a compensatory mechanism in which the reduction in mean arterial pressure caused by decreased total peripheral resistance triggers enhanced cardiac sympathetic activity, leading to augmented stroke volume and cardiac output. Previous studies of middle-aged and older adults have also reported that an impaired orthostatic increase in total peripheral resistance is primarily related to orthostatic hypotension [18, 19, 42].

Prior studies have established that impaired orthostatic increases in total peripheral resistance are linked to central neurodegenerative and peripheral autonomic disorders, including Parkinson’s disease and peripheral neuropathies [15, 43]. However, none of our participants had these diagnosed conditions. While diabetes mellitus is known to cause peripheral autonomic neuropathy, we found no significant association between diabetes mellitus and orthostatic decreases in TPRI. Arterial stiffness emerges as another potential mechanism underlying impaired orthostatic increases in total peripheral resistance. Research has demonstrated that arterial stiffness is associated with a smaller increase or a decrease in the mean arterial pressure upon standing [38] and orthostatic hypotension [44]. A previous head-up tilt study revealed that individuals exhibiting smaller TPRI increases and cardiac index decreases showed higher pulse wave velocity, indicating increased arterial stiffness [45, 46]. These individuals also demonstrated lower supine stroke and cardiac indices, higher supine TPRI, and a greater orthostatic increase in the ln(LF/HF) ratio; however, the causality remained undetermined [46]. Although we did not include pulse wave velocity measurements, the results of correlation analyses suggest that the observed orthostatic TPRI decrease is associated with arterial stiffness. Given that both stretching and aerobic exercises effectively reduce arterial stiffness [47, 48], future investigations should examine the effect of these exercise interventions on orthostatic changes in total peripheral resistance. Such research could clarify the mechanisms underlying impaired orthostatic total peripheral resistance increases and inform the development of targeted therapeutic strategies for orthostatic hypotension in stroke survivors.

### Study limitations

This study had some limitations. First, owing to an unexpected equipment malfunction, we were only able to include 23 participants instead of the planned 50 participants, which may have limited our statistical power. In addition, although orthostatic hypotension in stroke survivors is potentially associated with older age, female sex, hypertension, diabetes mellitus, low skeletal muscle mass, hypovolemia, severe motor paresis, and increased dependence in activities of daily living [12, 17], we did not observe associations of these characteristics with orthostatic hypotension or TPRI changes. This lack of association may be attributed to our limited sample size. Nevertheless, our results provide valuable insights into selecting the appropriate treatment for orthostatic hypotension based on individual hemodynamic patterns. Second, all participants were subacute stroke survivors receiving daily intensive rehabilitation, which may limit the generalizability of our findings to individuals with acute stroke who present with unstable general conditions or those in the chronic phase of stroke recovery after prolonged bed rest. Third, owing to the cross-sectional observational design of this study, we could not examine whether improvement in impaired orthostatic total peripheral resistance increases ameliorates orthostatic hypotension. Longitudinal studies are needed to investigate the temporal association between orthostatic hypotension and the orthostatic change in total peripheral resistance. Finally, this study did not include a control group. Further investigations should include an age- and sex-matched healthy control group for comparison with the stroke cohort. This approach would provide deeper insights into the pathogenesis of orthostatic hypotension and the impaired total peripheral resistance increase observed in individuals with stroke.

## Conclusions

In individuals with stroke, orthostatic hypotension was characterized by an impaired increase in total peripheral resistance rather than a marked decrease in cardiac output. These findings suggest that inadequate baroreflex-mediated vasoconstriction is the primary mechanism underlying orthostatic hypotension in this population. Our results enhance the understanding of hemodynamic responses to postural change after stroke and provide guidance for selecting appropriate therapeutic strategies.

## Acknowledgments

We would like to thank Editage (www.editage.com) for English language editing.

## Declaration of generative AI and AI-assisted technologies in the writing process

During the preparation of this work, the authors used Claude 3.5 Sonnet (Anthropic, San Francisco, CA, USA) for generating preliminary drafts and English editing assistance. After using this tool/service, the authors reviewed and edited the content as needed and took full responsibility for the content of the publication.

## Data availability statement

The data underlying this article will be shared on reasonable request to the corresponding author.

